# Lower State COVID-19 Deaths and Cases with Earlier School Closure in the U.S.

**DOI:** 10.1101/2020.05.09.20096594

**Authors:** Emily Rauscher

**Affiliations:** Department of Sociology, Brown University, Providence, RI 02912

## Abstract

This study quantifies the relationship between school closure timing and COVID-19 deaths and cases in the general population in all U.S. states. COVID-19 has higher symptomatic infection rates among the elderly, suggesting school closures could be unrelated to transmission. However, predicting daily cumulative COVID-19 deaths by state, earlier school closure is related to fewer deaths per capita and slower growth in deaths per capita. Results are similar for COVID-19 cases per capita.

## Introduction

Evidence is mixed on the extent to which spread of novel coronavirus or influenza strains slows with public school closure.^1,2^ Reviews find early school closures can slow influenza transmission when infection rates are higher among children than adults.^3,4^ Unlike most previous influenza outbreaks such as H1N1,^5^ COVID-19 causes higher symptomatic infection rates at older ages.^6^ School closures may therefore not slow the spread of COVID-19. To examine whether health benefits of closing schools outweigh the economic and social costs^5^, this study quantifies the relationship between state school closure timing and COVID-19 deaths and cases in the U.S.

## Methods

Using daily cumulative COVID-19 deaths and cases by state from *The New York Times*, this study examines dates after the first 100 cases in each state until April 27, 2020. COVID-19 prevalence is logged number of deaths (and cases) per 100,000 residents. Time period is days since the state reached 100 cases.

Using state-ordered school closure dates for each state from *EdWeek*, time to school closure is the number of days from when the state reached 100 cases until schools were closed. All states closed schools within 11 days after President Trump declared a national state of emergency on March 13. Variation in state prevalence rates on March 13 (zero to 6.6 cases/100,000) allows examination of the relationship between COVID-19 spread and school closure timing.

Equation 1 predicts log deaths per capita with time to school closure and the following controls to address potential confounding: 1) indicators for each state and date (to account for stable differences between states and national changes over time); and 2) state-level measures (represented by *X_it_* from 2020 World Population Data and *EdWeek)* of population (log); population density (log); number of public schools (log); public school enrollment (log); school closure date; stay-at-home order date (coded as last observed date for 5 states without stay-home orders); and an indicator for whether the state recommended rather than ordered school closure (sensitivity analyses exclude these 7 states and find similar results).

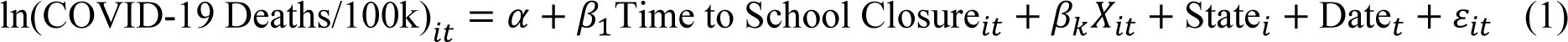

Growth in COVID-19 deaths is estimated by adding a control for lagged deaths (time *t-1*) in Equation 1. *β*_1_ then estimates the percent increase in deaths for each additional day from the time a state reached 100 cases until schools closed.

Data are weighted by the inverse of the number of days each state is observed to prevent states that reached 100 cases earlier from driving results. Robust standard errors are adjusted for state clustering.

## Results

Figure 1 shows cumulative state COVID-19 deaths per 100,000 residents by school closure timing. The bivariate correlation is 0.5 (p<0.001). Model 1 in Table 1 finds that deaths per 100,000 residents are predicted to be 12% higher for each additional day before schools closed (p<0.01). Controlling for lagged deaths (Model 3), state deaths increase by 1.3% for each additional day before schools closed (p<0.01). Estimates of both prevalence and growth are higher when including controls (Models 2 and 4).

**Figure 1:**
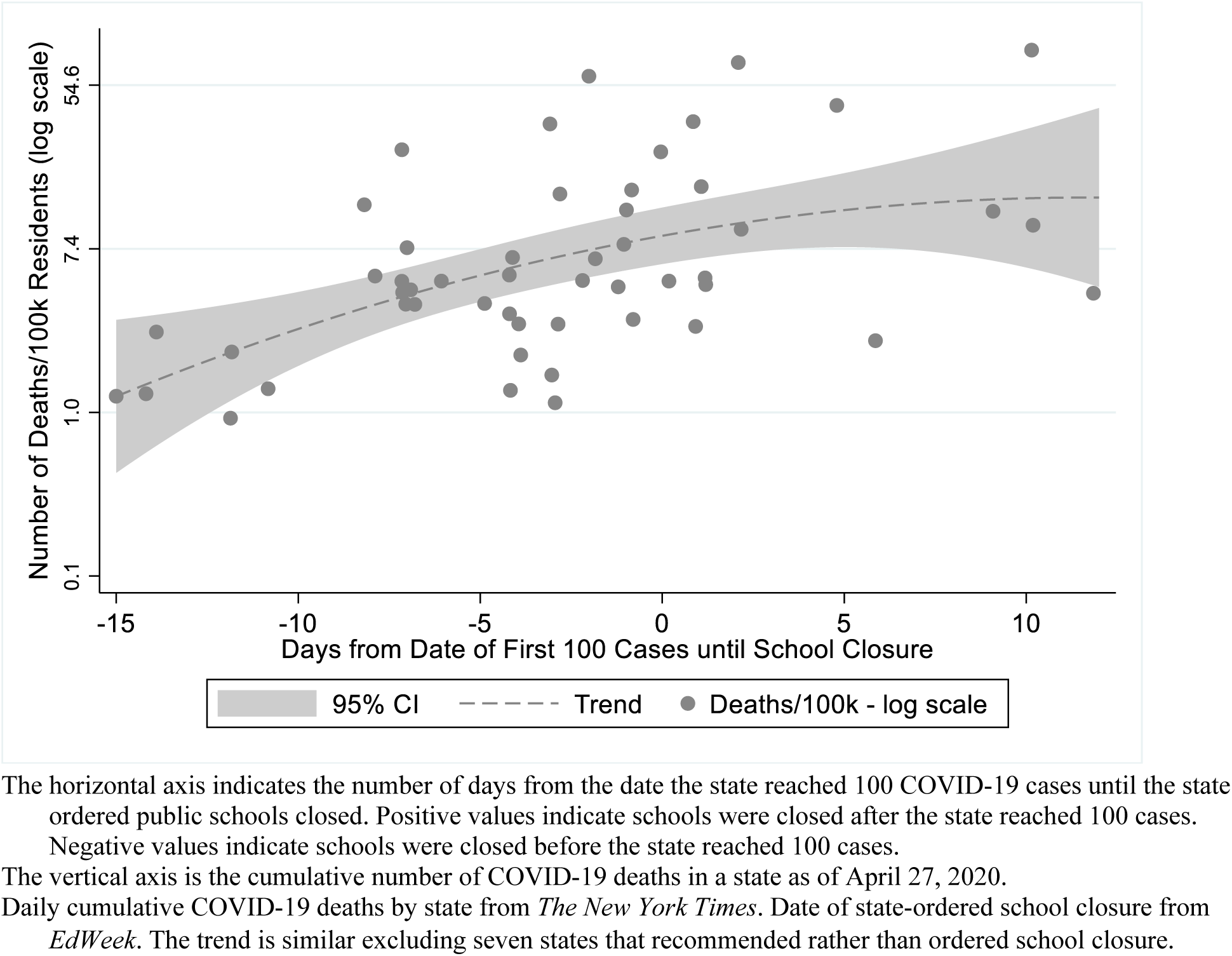
State COVID-19 Deaths by Timing of School Closure

**Table 1:**
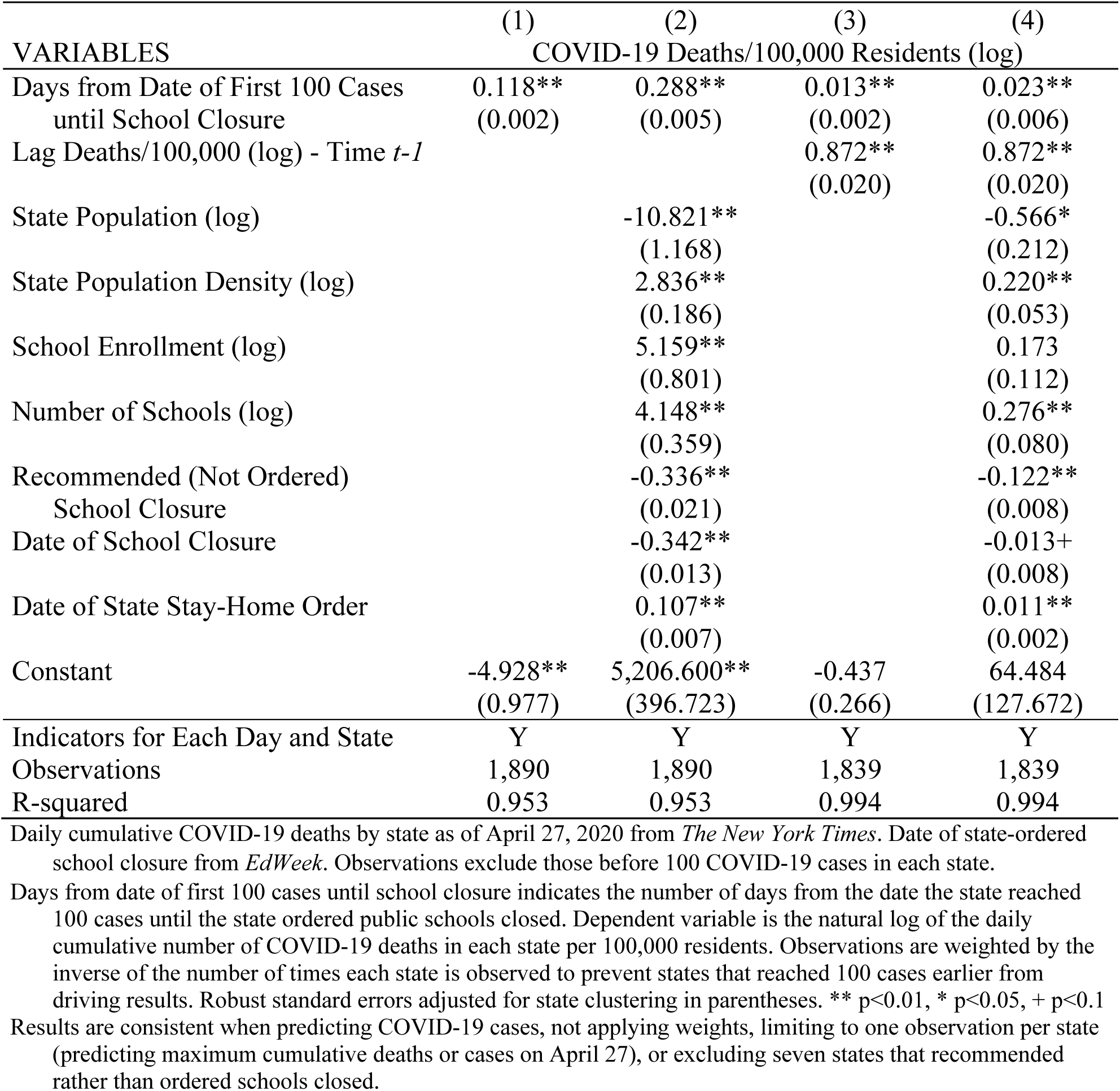
Predicted State COVID-19 Deaths per 100,000 Residents

| VARIABLES | (1) | (2) | (3) | (4) |
| --- | --- | --- | --- | --- |
|  | COVID-19 Deaths/100,000 Residents (log) |  |  |  |
| Days from Date of First 100 Cases<br>until School Closure | 0.118**<br>(0.002) | 0.288**<br>(0.005) | 0.013**<br>(0.002) | 0.023**<br>(0.006) |
| Lag Deaths/100,000 (log) - Time $t-1$ | | | 0.872**<br>(0.020) | 0.872**<br>(0.020) |
| State Population (log) |  | -10.821**<br>(1.168) |  | -0.566*<br>(0.212) |
| State Population Density (log) |  | 2.836**<br>(0.186) |  | 0.220**<br>(0.053) |
| School Enrollment (log) |  | 5.159**<br>(0.801) |  | 0.173<br>(0.112) |
| Number of Schools (log) |  | 4.148**<br>(0.359) |  | 0.276**<br>(0.080) |
| Recommended (Not Ordered)<br>School Closure |  | -0.336**<br>(0.021) |  | -0.122**<br>(0.008) |
| Date of School Closure |  | -0.342**<br>(0.013) |  | -0.013+<br>(0.008) |
| Date of State Stay-Home Order |  | 0.107**<br>(0.007) |  | 0.011**<br>(0.002) |
| Constant | -4.928**<br>(0.977) | 5,206.600**<br>(396.723) | -0.437<br>(0.266) | 64.484<br>(127.672) |
| Indicators for Each Day and State | Y | Y | Y | Y |
| Observations | 1,890 | 1,890 | 1,839 | 1,839 |
| R-squared | 0.953 | 0.953 | 0.994 | 0.994 |

Daily cumulative COVID-19 deaths by state as of April 27, 2020 from *The New York Times*. Date of state-ordered school closure from *EdWeek*. Observations exclude those before 100 COVID-19 cases in each state.
Days from date of first 100 cases until school closure indicates the number of days from the date the state reached 100 cases until the state ordered public schools closed. Dependent variable is the natural log of the daily cumulative number of COVID-19 deaths in each state per 100,000 residents. Observations are weighted by the inverse of the number of times each state is observed to prevent states that reached 100 cases earlier from driving results. Robust standard errors adjusted for state clustering in parentheses. \*\* $p < 0.01$ , \* $p < 0.05$ , + $p < 0.1$ . Results are consistent when predicting COVID-19 cases, not applying weights, limiting to one observation per state (predicting maximum cumulative deaths or cases on April 27), or excluding seven states that recommended rather than ordered schools closed.

Predicted cases per 100,000 are 2.7% higher and increase by 0.3% for each additional day before schools closed (p<0.01). Sensitivity analyses yield consistent results without applying weights, when limited to one observation per state (predicting maximum cumulative deaths or cases on April 27), or when excluding states that recommended rather than ordered schools closed.

## Discussion

This study quantifies the relationship between school closure timing and COVID-19 deaths and cases in the general population in all U.S. states. Although COVID-19 has higher symptomatic infection rates among the elderly^6^, earlier school closure is still related to fewer deaths per capita and slower growth in deaths per capita.

## Data Availability

All data are publicly available and code for data analysis is available upon request.

https://www.edweek.org/ew/section/multimedia/map-coronavirus-and-school-closures.html

https://worldpopulationreview.com/states/

https://github.com/nytimes/covid-19-data/blob/master/us-states.csv

https://www.wsj.com/articles/a-state-by-state-guide-to-coronavirus-lockdowns-11584749351

https://www.aljazeera.com/news/2020/03/emergencies-closures-states-handling-coronavirus-200317213356419.html

## Acknowledgments

This research was supported by the Spencer Foundation/National Academy of Education and by a grant from the American Educational Research Association which receives funds for its “AERA Grants Program” from the National Science Foundation under NSF Grant #DRL - 1749275. Opinions reflect those of the author and do not necessarily reflect those of the granting agencies.

## Appendix

**Table S1:**
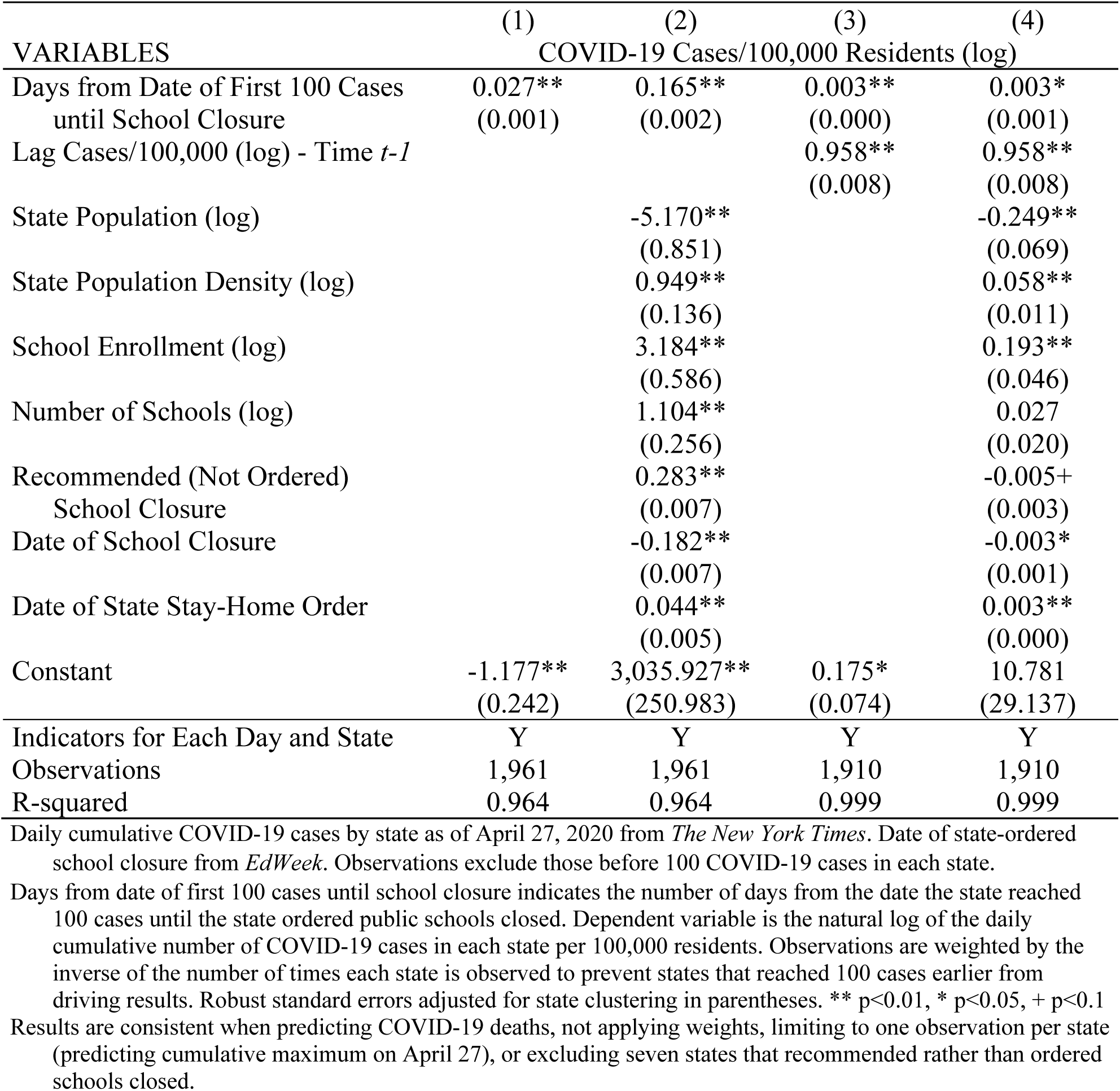
Predicted State COVID-19 Cases per 100,000 Residents

